# Extracorporeal Cytokine Hemadsorption in Severe COVID-19 Respiratory Failure

**DOI:** 10.1101/2020.06.28.20133561

**Authors:** Marianna Damiani, Lucia Gandini, Francesco Landi, Fabrizio Fabretti, Giuseppe Gritti, Ivano Riva

**Affiliations:** Dipartimento di Scienze della Salute, Università degli Studi di Milano, Milan, Italy; Intensive Care, Azienda Socio-Sanitaria Territoriale Papa Giovanni XXIII, Bergamo, Italy; Hematology Units, Azienda Socio-Sanitaria Territoriale Papa Giovanni XXIII, Bergamo, Italy

## Abstract

Despite the extracorporeal cytokine hemadsorption device CytoSorb was granted FDA emergency approval for critically ill COVID19 patients, to our knowledge no published studies are currently available to support its use. This manuscript reports the experience of the use of CytoSorb during COVID19 pandemic in Bergamo, Italy. In our pilot study, eleven COVID19 patients requiring invasive mechanical ventilation for a rapidly progressive ARDS were treated with 24 to 48 hours of extracorporeal cytokine hemadsorption.

Respiratory and laboratory parameters, including a full set of inflammatory cytokines, were evaluated at different time points. A significant but transient reduction of the hyperinflammatory status was observed, along with the amelioration of the clinical and respiratory parameters.

We believe that this manuscript will provide them with important preliminary data on the use of cytokine hemadsorption devices.

## To the Editor

Nearly 20% of patients infected by severe acute respiratory syndrome coronavirus-2 (SARS-CoV-2) experience a severe disease requiring admission to intensive care unit (ICU) with mortality rates up to 26% of the cases.^1 2^Hyper-inflammatory activation characterized by immune cell infiltration and elevated levels of cytokines as interleukin (IL)-6, IL-8 and C-reactive protein (CRP) was reported as the main mechanism leading to critical illness and severe acute respiratory distress syndrome (ARDS).^3^ Thus, targeting the cytokine storm may represent a promising therapeutic strategy. While the use of monoclonal antibodies targeting the IL-6 pathway was reported to improve outcome, a significant increase of bacterial infections has been observed.^4^

An alternative treatment able to reduce circulating cytokines in critically ill patients is CytoSorb (Aferetica srl, Italy), a device containing adsorbent polymer beads designed to irreversibly remove cytokines currently used for septic shock and other conditions where elevated levels of cytokines are present.^5^ Along with the beneficial effect on system inflammation, CytoSorb can be easily integrated with all extracorporeal circulation systems, as continuous renal replacement therapy (CRRT) or ExtraCorporeal Membrane Oxygenation (ECMO). Despite the lack of published results, based on bench performance testing and reported clinical experience, CytoSorb was granted FDA emergency approval for critically ill SARS-CoV-2 patients on April 10, 2020.^6^

In this report we present the laboratory and clinical outcome of Corona Virus Disease 2019 (COVID19) patients treated with CytoSorb at Papa Giovanni XXIII Hospital, Bergamo, Italy. The study was approved by the Ethic Committee and patients, or their legal representative, provided either verbal or written consent to participate in the study. Clinical data were gathered from the electronic charts. Inflammatory cytokines including IL-1β, IL-6, IL-8, IL-10 IL-12p70 and tumor necrosis factor (TNF)-α were dosed in a single serum sample by flow cytometry (BD CBA Human Inflammatory Cytokines Kit, BD Biosciences, USA). Non-parametric multiple comparison Kurskalis-Wallis tests were performed to analyse cytokines levels evolution over time. Descriptive statistics and all tests were performed using NCSS 10 (Statistical Software 2015 NCSS, LLC. Kaysville, Utah, USA).

From March 7 to March 24, 2020, 11 patients with microbiological confirmed SARS-CoV-2 infection and severe disease requiring invasive mechanical ventilation (IMV) were treated with CytoSorb. A Kimall catheter was inserted and CRRT was started with continuous veno-venous hemodiafiltration (CVVHDF) modality and high ultrafiltration rate. The adsorber was changed every 24 hours to avoid cytokines release from cartridge saturation. Two patients were treated with one 24-hour cycle, while the remaining 9 were treated for 48 hours. Best supportive care was provided according to hospital guidelines and included antiviral therapy (lopinavir/ritonavir 200/50 mg two tablets twice daily, darunavir/cobicistat 800/150 mg one tablet once daily) and hydroxychloroquine 200 mg twice daily.

Table 1 summarized patient’s baseline clinical characteristics. All patients were male, overweight and only 3 (27%) were over 70 years old. Despite none of them had severe comorbidity, patients presented an aggressive and rapidly evolutive disease with need non-invasive ventilation in all of the cases followed by IMV after a median of 3 days (range 0-4 days) from hospital admission.

**Table 1.** Baseline patient and disease characteristics BMI: body mass index; SOFA: Sequential Organ Failure Assessment; SAPS: Simplified Acute Physiology Score; ICU: intensive care unit; CRP: C-reactive protein.

|  | **N (%) or median (range)** |
| --- | --- |
| **Male sex** | 11 (100%) |
| **Age** (years) | 62 (35–75) |
| **BMI** (kg/m<sup>2</sup>) | 28 (22–37) |
| **Comorbidities** |  |
| *Hypertension* | 7 (64%) |
| *Diabetes* | 3 (27%) |
| *Cardiovascular disease* | 0 (0%) |
| *Malignancies* | 0 (0%) |
| *Cerebrovascular disease* | 2 (18%) |
| *Chronic kidney disease* | 0 (0%) |
| **Enrollment from intubation** (days) | 3 (0–8) |
| **SOFA score** | 4 (4.5–6.5) |
| **SAPS II score** | 40 (29–56) |
| **PaO<sub>2</sub>/FiO<sub>2</sub>** (ratio) | 103 (84–180) |
| **CRP** (mg/dl) | 29 (4–42) |

Cytokines levels were evaluated before and immediately after treatment. In addition, samples were collected after 24 hours and 7 days from treatment end. Before treatment, a markedly elevated levels of IL-6 and IL-8 were observed, with median values of 355 pg/mL (interquartile range, IQR 263-466) and 107 pg/mL (IQR 46-198), respectively. The other cytokines, including IL-1β, IL-10, IL-12p70 and TNF-α, were not significantly elevated. A significant reduction of IL-6 compared to baseline was observed at treatment end and 24 hours after, with median levels of 118 pg/mL (IQR 19-221, p=0,003) and 169 pg/mL (IQR 61-253, p=0,03), respectively. Similarly, a non-significant trend toward reduction was observed for IL-8 at the same time point compared to baseline, with median levels of 55 pg/mL (IQR 45-76, p=0,16) and 67 pg/mL (IQR 41-86, p=0,36), respectively. However, after 7 days from the end of the treatment an increase of both IL-6 and IL-8 was noticed, with median values of 159 pg/mL (IQR 107-419, p=0,22) and 77 pg/mL (IQR 54-200, p=0,21), respectively.

C-reactive protein (CRP) levels and respiratory parameters including the PaO2/FiO2 ratio (P/F) were assessed daily. Consistent with the trend observed for IL-6, a significant drop of CRP median levels was observed starting from 2 days after treatment and for the subsequent 4 days, followed by a new increase (Figure 1). Median P/F value at enrolment was 103 (IQR 88-133), consistent with a moderate to severe ARDS. The decrease in the inflammatory status was associated with a progressive improvement in the respiratory function, with a significant increase in P/F from the first day after end of the therapy (Figure 1).

**Figure 1.**
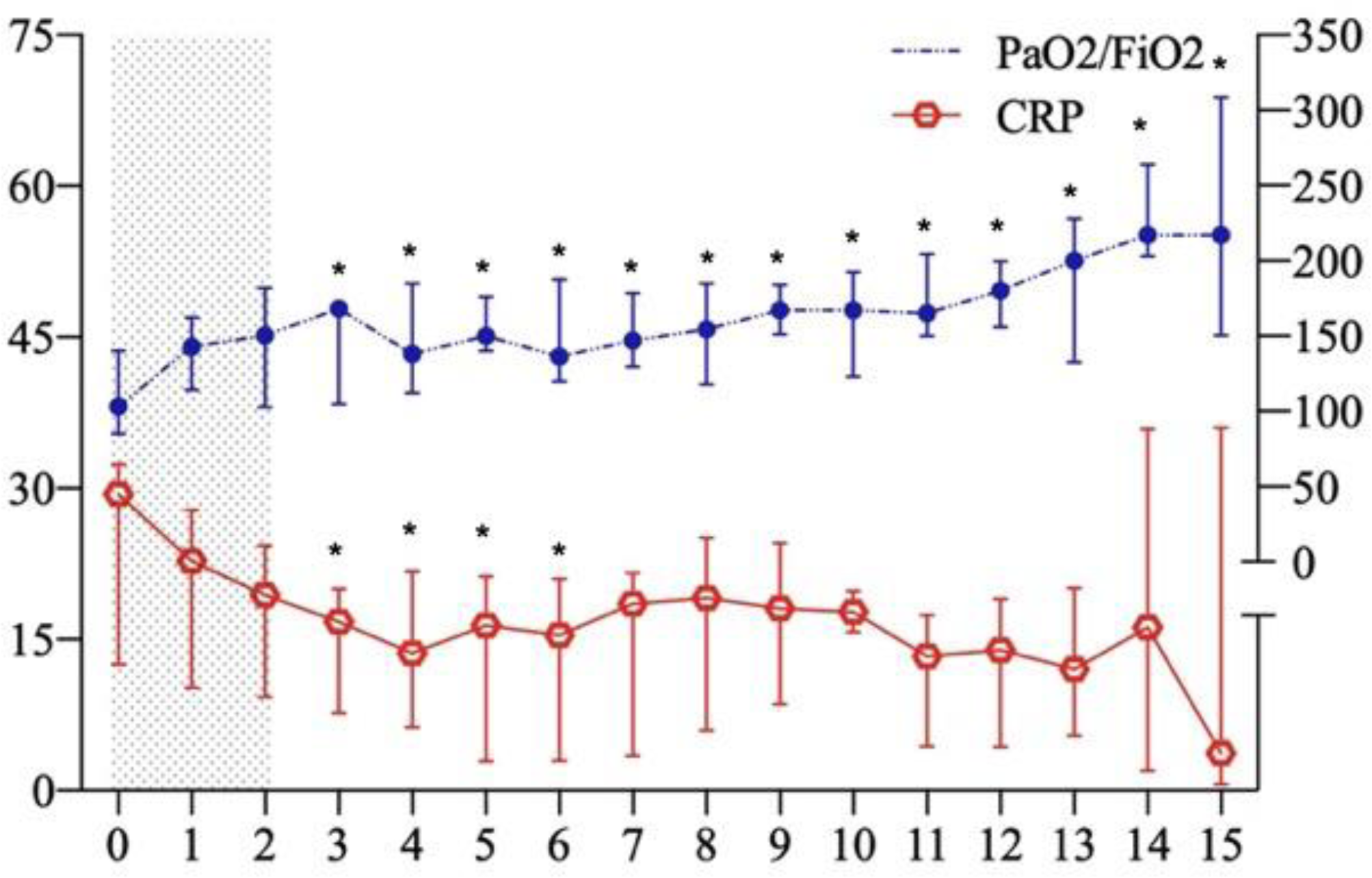
CRP and PaO_2_/FiO_2_ ratio trends (median, IQR) during first 15 days from enrollment. Dotted area represents treatment. CRP: C-reactive protein (CRP), IQR: interquartile ranges. *P <0.05 referred to baseline

No unexpected adverse events were observed with CytoSorb use. After a median follow-up of 16 days (range 6-30 days), 2 patients died (18%) after 6 and 16 days from enrollment, and 9 are still alive. In 3 patients weaning was completed with extubation at day 14, 15 and 27 from enrollment. Six patients required percutaneous tracheostomy to achieve respiratory weaning. All 9 patients were discharged from ICU after a median of 25 days (range 11-52).

In this study we report for the first time that CytoSorb treatment for 24-48 hours significantly reduces IL-6 levels resulting in a beneficial effect on systemic inflammation in the subsequent days. However, the persistence of significant levels of IL-6 and CRP after treatment and their increase at one week suggest that a more prolonged treatment is required to control SARS-CoV-2 hyperinflammatory status. Since no specific guidelines were available at the time, CytoSorb was used empirically based on previous experience in septic shock. However, the results support the proposed CytoSorb use with 72 hours long treatment with 4 adsorbers. With the limitation of the small sample size, CytoSorb proved to be safe and a clinical amelioration was observed in most of the treated patients despite the severity of the disease. Further studies are required to assess if CytoSorb can improve the clinical outcome of critically ill patients.

## Data Availability

Descriptive statistics and all tests were performed using NCSS 10 (Statistical Software 2015 NCSS, LLC. Kaysville, Utah, USA).

## Acknowledgments

This work was supported in part by an unrestricted grant from Aferetica srl, Italy. We would like to thank Dr. Andjela Kurevija (Aferetica srl) for the statistical support.

## Notes

**Declaration of interests:** GG reports non-financial support from Gilead Kite, Roche, Takeda and Janssen, and personal fees from Gilead Kite, Autolus, Roche, IQvia, Takeda, Amgen and Italfarmaco, outside the submitted work. Ivano Riva reports travel support from Aferetica. All other authors declare no competing interests.

### Competing Interest Statement

Declaration of interests: GG reports non-financial support from Gilead Kite, Roche, Takeda and Janssen, and personal fees from Gilead Kite, Autolus, Roche, IQvia, Takeda, Amgen and Italfarmaco, outside the submitted work. Ivano Riva reports travel support from Aferetica. All other authors declare no competing interests.

### Author Declarations

The study was approved by the Ethic Committee (ASST Papa Giovanni XXIII, resolution n 257/2020) and patients, or their legal representative, provided either verbal or written consent to participate in the study. ethics committee

